# Development of a Liver Disease-Specific Large Language Model Chat Interface using Retrieval Augmented Generation

**DOI:** 10.1101/2023.11.10.23298364

**Authors:** Jin Ge, Steve Sun, Joseph Owens, Victor Galvez, Oksana Gologorskaya, Jennifer C. Lai, Mark J. Pletcher, Ki Lai

**Author notes:** Correspondence: Jin Ge, MD, MBA, 513 Parnassus Avenue, S-357, San Francisco, CA 94143.

## Abstract

**Background:** Large language models (LLMs) have significant capabilities in clinical information processing tasks. Commercially available LLMs, however, are not optimized for clinical uses and are prone to generating incorrect or hallucinatory information. Retrieval-augmented generation (RAG) is an enterprise architecture that allows embedding of customized data into LLMs. This approach “specializes” the LLMs and is thought to reduce hallucinations.

**Methods:** We developed “LiVersa,” a liver disease-specific LLM, by using our institution’s protected health information (PHI)-complaint text embedding and LLM platform, “Versa.” We conducted RAG on 30 publicly available American Association for the Study of Liver Diseases (AASLD) guidelines and guidance documents to be incorporated into LiVersa. We evaluated LiVersa’s performance by comparing its responses versus those of trainees from a previously published knowledge assessment study regarding hepatitis B (HBV) treatment and hepatocellular carcinoma (HCC) surveillance.

**Results:** LiVersa answered all 10 questions correctly when forced to provide a “yes” or “no” answer. Full detailed responses with justifications and rationales, however, were not completely correct for three of the questions.

**Discussions:** In this study, we demonstrated the ability to build disease-specific and PHI-compliant LLMs using RAG. While our LLM, LiVersa, demonstrated more specificity in answering questions related to clinical hepatology – there were some knowledge deficiencies due to limitations set by the number and types of documents used for RAG. The LiVersa prototype, however, is a proof of concept for utilizing RAG to customize LLMs for clinical uses and a potential strategy to realize personalized medicine in the future.

## Introduction

Large language models (LLMs), such as OpenAI’s Generative Pre-trained Transformer (GPT) family of models, have demonstrated significant capabilities in clinical information processing tasks, such as data extraction,^1^ summarizing literature,^2^ content generation,^3^ and predictive modeling.^4^ Commercially available general-purpose LLMs, such as OpenAI’s ChatGPT, however, are trained on publicly available data and not optimized for clinical uses.^5^ This means that the outputs of publicly available LLMs when prompted with clinical questions may include incorrect, incomplete, or hallucinatory information.^6,7^ Despite these limitations, LLMs are thought to have significant potential in biomedical and clinical applications. This is because the practice of modern medicine is highly complex endeavor with ever-increasing amounts of knowledge generated yearly.^8^ For instance, it was estimated that two papers were uploaded to PubMed every minute in 2016,^9^ a figure that has undoubtedly increased in the seven years since. National practice societies have also responded to the ever-expanding body of clinical knowledge by developing comprehensive practice guidelines. For instance, the American Association of the Study of Liver Diseases (AASLD) has issued guidelines and guidance documents for 26 liver diseases and conditions, and two quality measures concerning cirrhosis and hepatocellular carcinoma (HCC).^10^ Methods to incorporate medical literature, such as clinical practice guidelines and guidance documents, into LLM responses, therefore, represent growing area of interest to help general-purpose LLMs become “specialized” for clinically-focused applications.

Currently, there are three general approaches and techniques to allow LLM “specialization:” 1. Fine-tune the original LLM model, which is computationally expensive;^11,12^ 2. Prompting within the LLM, which only accommodates small amounts of data and requires iterative user input;^13–15^ and 3. Retrieval-augmented generation (RAG).^16–18^ RAG is an enterprise architecture that augments LLM abilities by adding an information retrieval system that provides external data, which then supplement and constrain LLM output. In practice, this means that a dataset, such as a compendium of clinical practice guidelines, is vectorized and encoded using embedding models and then incorporated into the LLM by layering it on top of the LLM information retrieval and output processes.^16^ The theoretical advantages of RAG are two-fold: 1. Handling large numbers of documents to provide “ground knowledge” for the LLM, and 2. Decreasing hallucinations by limiting the potential “solution-space” for LLM outputs. This RAG approach has been proposed and utilized in other clinical specialties, such as general medicine and urology.^17,19,20^ Existing RAG implementations, however, have largely been on publicly available and not protected health information [PHI]-compliant LLMs – thereby limiting permitted use in clinical practice

In this study, we utilized RAG to create a prototype liver disease-specific LLM, called “LiVersa,” within the University of California, San Francisco’s (UCSF) PHI-compliant implementation of Microsoft OpenAI GPT family of LLM models, “Versa.”

## Methods

This study was not considered human subjects research because no human data or specimens were used. Consequently, approval from the UCSF Institutional Review Board was not sought. To construct our liver disease-specific LLM (LiVersa), we used all available provider-facing clinical practice guidelines, guidance documents, and quality measure documents published by AASLD on its website.^10^ We excluded introduction and executive summary documents as the content in these were often duplicated in the corresponding full-length versions (e.g., for Wilson’s Disease). We included both original guidelines/guidance documents and updates if both were publicly available for download on AASLD website (e.g., for Chronic Hepatitis B [HBV]). Patient-facing and/or outdated guidelines/guidance documents were not included in this study. In total, 30 documents were retrieved to be incorporated into the RAG process (Table 1).

**Table 1.** AASLD Guidelines, Guidance Documents, and Quality Measure Documents Incorporated in LiVersa.

| # | Journal | Year | Title |
| --- | --- | --- | --- |
| 1 | Hepatology | 2011 | AASLD Position Paper: The Management of Acute Liver Failure: Update 2011<sup>32</sup> |
| 2 | Hepatology | 2020 | Diagnosis and Treatment of Alcohol-Associated Liver Diseases: 2019 Practice Guidance From the American Association for the Study of Liver Diseases<sup>33</sup> |
| 3 | Hepatology | 2021 | Diagnosis, Evaluation, and Management of Ascites, Spontaneous Bacterial Peritonitis and Hepatorenal Syndrome: 2021 Practice Guidance by the American Association for the Study of Liver Diseases<sup>34</sup> |
| 4 | Hepatology | 2020 | Diagnosis and Management of Autoimmune Hepatitis in Adults and Children: 2019 Practice Guidance and Guidelines From the American Association for the Study of Liver Diseases<sup>35</sup> |
| 5 | Hepatology | 2023 | AASLD practice guidance on drug, herbal, and dietary supplement–induced liver injury<sup>36</sup> |
| 6 | Hepatology | 2011 | Diagnosis and management of hemochromatosis: 2011 Practice Guideline by the American Association for the Study of Liver Diseases<sup>37</sup> |
| 7 | Hepatology | 2014 | Hepatic encephalopathy in chronic liver disease: 2014 Practice Guideline by the American Association for the Study Of Liver Diseases and the European Association for the Study of the Liver<sup>38</sup> |
| 8 | Hepatology | 2016 | AASLD guidelines for treatment of chronic hepatitis B<sup>39</sup> |
| 9 | Hepatology | 2018 | Update on prevention, diagnosis, and treatment of chronic hepatitis B: AASLD 2018 hepatitis B guidance<sup>40</sup> |
| 10 | Clinical Infectious Diseases | 2023 | Hepatitis C Guidance 2023 Update: American Association for the Study of Liver Diseases–Infectious Diseases Society of America Recommendations for Testing, Managing, and Treating Hepatitis C Virus Infection<sup>41</sup> |
| 11 | Hepatology | 2023 | AASLD Practice Guidance on prevention, diagnosis, and treatment of hepatocellular carcinoma<sup>27</sup> |
| 12 | Hepatology | 2009 | AASLD Position Paper: Liver biopsy<sup>42</sup> |
| 13 | Hepatology | 2014 | Evaluation for liver transplantation in adults: 2013 practice guideline by the American Association for the Study of Liver Diseases and the American Society of Transplantation<sup>43</sup> |
| 14 | Hepatology | 2014 | Evaluation of the Pediatric Patient for Liver Transplantation: 2014 Practice Guideline by the American Association for the Study of Liver Diseases, American Society of Transplantation and the North American Society for Pediatric Gastroenterology, Hepatology and Nutrition<sup>44</sup> |
| 15 | Liver Transplantation | 2013 | Long-Term Management of the Successful Adult Liver Transplant: 2012 Practice Guideline by the American Association for the Study of Liver Diseases and the American Society of Transplantation<sup>45</sup> |
| 16 | Liver Transplantation | 2013 | Long-term medical management of the pediatric patient after liver transplantation: 2013 practice guideline by the American Association for the Study of Liver Diseases and the American Society of Transplantation<sup>46</sup> |
| 17 | Hepatology | 2021 | Malnutrition, Frailty, and Sarcopenia in Patients With Cirrhosis: 2021 Practice Guidance by the American Association for the Study of Liver Diseases<sup>47</sup> |
| 18 | Hepatology | 2023 | AASLD Practice Guidance on the clinical assessment and management of nonalcoholic fatty liver disease<sup>48</sup> |
| 19 | Hepatology | 2022 | AASLD Practice Guidance: Palliative care and symptom-based management in decompensated cirrhosis<sup>49</sup> |
| 20 | Hepatology | 2023 | AASLD practice guidance on risk stratification and management of portal hypertension and varices in cirrhosis<sup>50</sup> |
| 21 | Hepatology | 2023 | AASLD Practice Guidance on the Use of TIPS, Variceal Embolization, and Retrograde Transvenous Obliteration in the Management of Variceal Hemorrhage<sup>51</sup> |
| 22 | Hepatology | 2019 | Primary Biliary Cholangitis: 2018 Practice Guidance from the American Association for the Study of Liver Diseases<sup>52</sup> |
| 23 | Hepatology | 2022 | Primary biliary cholangitis: 2021 practice guidance update from the American Association for the Study of Liver Diseases<sup>53</sup> |
| **24** | Hepatology | 2023 | AASLD practice guidance on primary sclerosing cholangitis and cholangiocarcinoma<sup>54</sup> |
| **25** | Hepatology | 2021 | Reproductive Health and Liver Disease: Practice Guidance by the American Association for the Study of Liver Diseases<sup>55</sup> |
| **26** | Hepatology | 2021 | Vascular Liver Disorders, Portal Vein Thrombosis, and Procedural Bleeding in Patients With Liver Disease: 2020 Practice Guidance by the American Association for the Study of Liver Diseases<sup>56</sup> |
| **27** | Hepatology | 2022 | A multidisciplinary approach to the diagnosis and management of Wilson disease: 2022 Practice Guidance on Wilson disease from the American Association for the Study of Liver Diseases<sup>57</sup> |
| **28** | Hepatology | 2018 | Development of Quality Measures in Cirrhosis by the Practice Metrics Committee of the American Association for the Study of Liver Diseases<sup>58</sup> |
| **29** | Hepatology | 2021 | Quality measures in HCC care by the Practice Metrics Committee of the American Association for the Study of Liver Diseases<sup>59</sup> |
| **30** | Hepatology | 2023 | AASLD Practice Guidance on Acute-on-chronic Liver Failure and the Management of Critically Ill Patients with Cirrhosis<sup>60</sup> |

We utilized application programming interfaces (APIs) provided by the Microsoft Azure OpenAI Cognitive Search suite of tools to incorporate the AASLD guidelines and documents for RAG (Figure 1).^16^ In the pre-processing phase, the 30 AASLD guidelines and documents in PDF format were transformed into embeddings using Microsoft Azure OpenAI’s ADA Text Embedding Version 2 model (*text-embedding-ada-002*). Text embeddings are numerical representations of text where words for phrases are represented as multi-dimensional vectors.^21,22^ These embedding vectors are then stored in a database with the Microsoft Azure Cognitive Search services. During chat interactions with LiVersa, users’ prompts are converted into embeddings in real time using the same *text-embedding-ada-002* model. A search is then performed on the previously processed vector database of AASLD guidelines and documents to find matches for the prompt embeddings. The search results from this search process are then passed to the *gpt-35-turbo or gpt-4-32k LLMs* to generate a completion, which is the output from the LLM in the LiVersa chat interface.

**Figure 1.**
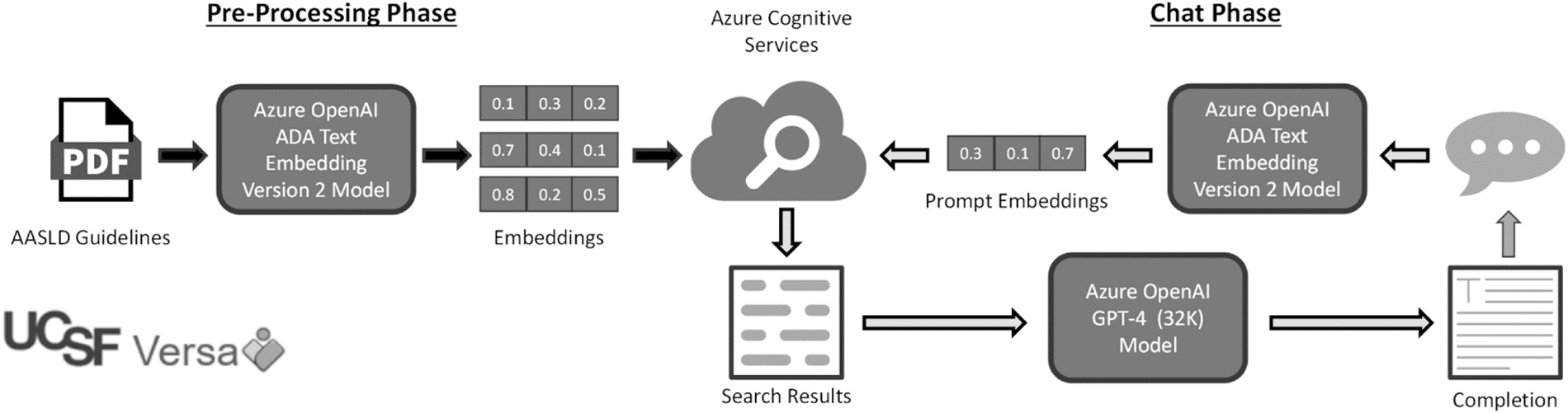
Diagram for Retrieval Augmented Generated as Executed Through Azure Cognitive Search.

We pre-set three sample questions into “Sample Questions” section of the LiVersa interface:

1. What are the indications for liver transplantation?
2. Who should be screened for chronic hepatitis B?
3. What is the recommended therapy for a patient with BCLC 0-A HCC without portal hypertension?

To compare LiVersa’s performance on free-form clinical hepatology questions, we compared its responses to medical trainees’ responses in a previously published case-vignette based knowledge assessment questions on HBV treatment and hepatocellular carcinoma (HCC) surveillance.^23^ We chose this set of questions because they have also been evaluated by the publicly available ChatGPT.^24^ Prior to input into LiVersa, case-vignettes were edited for clarity and appropriateness with the goal of retaining the essence of the clinical scenario. We asked LiVersa to give both a full response and a forced “yes” or “no” response by appending the prompt with “Answer yes or no” to each inputted case-vignette (Figure 2).

**Figure 2.**
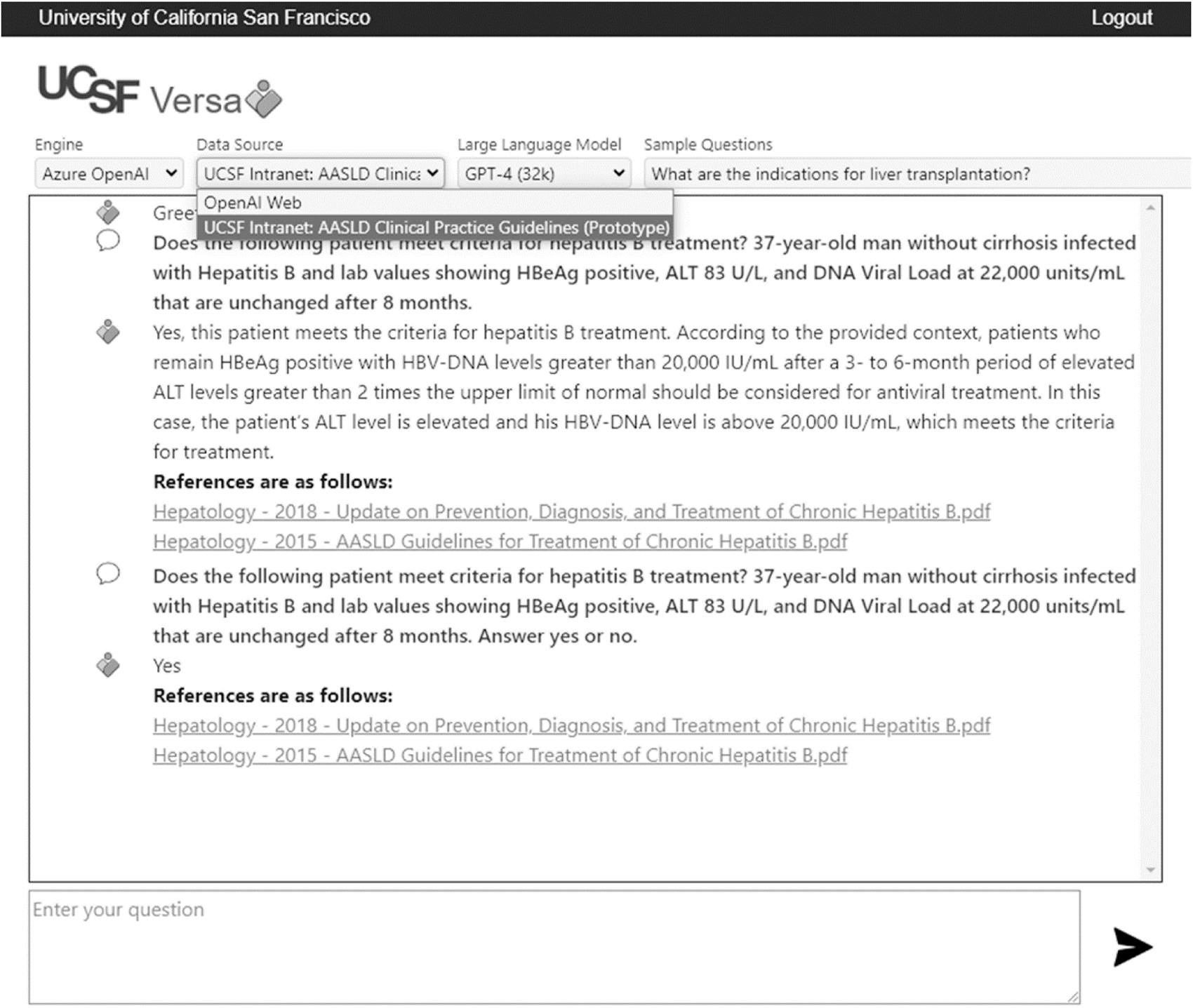
LiVersa Chat Interface.

## Results

The user interface for LiVersa is shown in Figure 2. The interface is the same as to the one used for “Versa Chat” (the chat interface for the general-purpose PHI-compliant LLM implementation at UCSF), except with the data source being set to the “UCSF Intranet: AASLD Clinical Practice Guidelines (Prototype).” LiVersa’s responses to the three sample questions are featured in Table 2. LiVersa’s performance on case-vignette based knowledge assessment questions on HBV treatment and HCC surveillance are featured in Table 3, Columns B and C. The correct answers along with percentages of trainees who answered correctly per prior studies are featured in Table 3, Column D. LiVersa answered all 10 questions correctly when forced to provide a “yes” or “no” answer. Rationales and justifications within the full answers, however, were not completely correct for three clinical scenarios in HCC surveillance. These three clinical scenarios were: “A 25-year-old Haitian man with chronic hepatitis B, on treatment with entecavir,” “A 40-year-old Cuban woman who was recently diagnosed with hepatitis B after developing jaundice in the setting of a surgical procedure. There is no evidence of cirrhosis,” and “A 40-year-old woman from Thailand with cirrhosis and chronic inactive hepatitis B.”

**Table 2.** LiVersa Responses to “Sample Questions” Embedded in the Chat Interface.

| Sample Question | LiVersa Generated Answer |
| --- | --- |
| ***What are the indications for liver transplantation?*** | The indications for liver transplantation include severe acute or advanced chronic liver disease when the limits of medical therapy have been reached. Specific conditions that may necessitate a liver transplant include:<br>- Acute Liver Failure<br>- Complications of cirrhosis such as ascites, chronic gastrointestinal blood loss due to portal hypertensive gastropathy, encephalopathy, refractory variceal hemorrhage, and synthetic dysfunction<br>- Liver cancer<br>- Liver-based metabolic conditions with systemic manifestations such as alpha-1 antitrypsin deficiency, familial amyloidosis, glycogen storage disease, hemochromatosis, primary oxaluria, and Wilson disease<br>- Systemic complications of chronic liver disease such as hepatopulmonary syndrome and portopulmonary hypertension<br>Evaluation for liver transplantation should be considered once a patient with cirrhosis has experienced an index complication such as ascites, hepatic encephalopathy, or variceal hemorrhage or hepatocellular dysfunction results in a MELD Score > 15. Potential liver transplant candidates with worsening renal dysfunction or other evidence of rapid hepatic decompensation should have prompt evaluation for liver transplant. |
| ***Who should be screened for chronic hepatitis B?*** | Screening for chronic Hepatitis B is recommended for the following groups:<br>1. Persons born in regions of high or intermediate HBV endemicity (HBsAg prevalence of $\geq 2\%$)<br>2. U.S.-born persons not vaccinated as an infant whose parents were born in regions with high HBV endemicity ($\geq 8\%$)<br>3. Persons who have ever injected drugs<br>4. Men who have sex with men<br>5. Persons needing immunosuppressive therapy, including chemotherapy, immunosuppression related to organ transplantation, and immunosuppression for rheumatological or gastroenterologic disorders<br>6. Individuals with elevated ALT or AST of unknown etiology<br>7. Donors of blood, plasma, organs, tissues, or semen<br>8. Persons with end-stage renal disease, including predialysis, hemodialysis, peritoneal dialysis, and home dialysis patients<br>9. All pregnant women<br>10. Infants born to HBsAg-positive mothers<br>11. Persons with chronic liver disease, e.g., HCV<br>12. Persons with HIV<br>13. Household, needle-sharing, and sexual contacts of HBsAg-positive persons<br>14. Persons who are not in a long-term, mutually monogamous relationship (e.g., >1 sex partner during the previous 6 months)<br>15. Persons seeking evaluation or treatment for a sexually transmitted disease<br>16. Health care and public safety workers at risk for occupational exposure to blood or blood-contaminated body fluids<br>17. Residents and staff of facilities for developmentally disabled persons<br>18. Travelers to countries with intermediate or high prevalence of HBV infection<br>19. Persons who are the source of blood or body fluid exposures that might require postexposure prophylaxis<br>20. Inmates of correctional facilities<br>21. Unvaccinated persons with diabetes who are aged 19 through 59 years (discretion of clinician for unvaccinated adults with diabetes who are aged $\geq 60$ years). |
| ***What is the recommended therapy for a patient with BCLC 0-A HCC without portal hypertension?*** | The expert panel recommends that surgical resection should be considered in relevant cases for patients with BCLC 0-A HCC without portal hypertension. |

**Table 3.** LiVersa Responses to HBV Treatment and HCC Surveillance Questions.

| Case Vignette (Edited for Clarity and Appropriateness) | LiVersa Full Response | LiVersa Yes/No Response | Correct Answer And %Correct Amongst Trainees<sup>23</sup> |
| --- | --- | --- | --- |
| ***Does the following patient meet criteria for hepatitis B treatment?*** |  |  |  |
| ***37-year-old man without cirrhosis infected with Hepatitis B and lab values showing HBeAg positive, ALT 83 U/L, and DNA Viral Load at 22,000 units/mL that are unchanged after 8 months.*** | Yes, this patient meets the criteria for hepatitis B treatment. According to the provided context, patients who remain HBeAg positive with HBV-DNA levels greater than 20,000 IU/mL after a 3- to 6-month period of elevated ALT levels greater than 2 times the upper limit of normal should be considered for antiviral treatment. In this case, the patient's ALT level is elevated and his HBV-DNA level is above 20,000 IU/mL, which meets the criteria for treatment. | Yes | Yes - 78.9% |
| ***55-year-old woman with compensated cirrhosis secondary to Hepatitis B with lab values showing HBeAg negative, DNA Viral load at 4,000 units/mL and ALT 35 U/L.*** | Yes, this patient meets the criteria for hepatitis B treatment. According to the guidelines, therapy is recommended for persons with immune-active chronic hepatitis B (CHB) and cirrhosis if HBV DNA is >2,000 IU/mL, regardless of ALT level. This patient's HBV DNA level is 4,000 IU/mL, which is above the threshold, and she has cirrhosis. Therefore, she should be considered for antiviral therapy. | Yes | Yes - 49.2% |
| ***32-year-old man without cirrhosis infected with Hepatitis B with lab values showing HBeAg positive, DNA Viral Load at 8000 units/mL and ALT 22 U/L.*** | Based on the provided guidelines, this patient does not meet the criteria for treatment. The guidelines recommend antiviral therapy for adults with immune-active chronic hepatitis B (CHB), which is defined by an elevation of ALT more than 2 times the upper limit of normal (ULN) or evidence of significant histologic disease plus elevated HBV DNA above 2,000 IU/mL (HBeAg negative) or above 20,000 IU/mL (HBeAg positive). This patient's ALT level is within the normal range and his HBV DNA level is below the threshold for HBeAg positive patients. Therefore, he does not meet the criteria for treatment. However, regular monitoring of disease status is recommended to determine the need for antiviral therapy. | No | No - 22.2% |
| ***Should the following patient be screened for hepatocellular carcinoma?*** |  |  |  |
| ***A 25-year-old Haitian man with chronic hepatitis B, on treatment with entecavir.\**** | According to the provided context, the patient should be screened for hepatocellular carcinoma. The context states that non-cirrhotic chronic hepatitis B patients, especially men from endemic countries over the age of 40, should be screened for hepatocellular carcinoma. Although the patient is younger than 40, he is from an endemic country and has chronic hepatitis B, which puts him at risk. | Yes | Yes - 71.1% |
| ***A 40-year-old Cuban woman who was recently diagnosed with hepatitis B after developing jaundice in the setting of a surgical procedure. There is no evidence of cirrhosis.\**** | Based on the provided context, this patient does not meet the criteria for hepatocellular carcinoma (HCC) surveillance. The guidelines suggest that non-cirrhotic chronic hepatitis B patients who are men from endemic countries over the age of 40 or women from endemic countries over the age of 50 should be screened. However, this patient is a 40-year-old woman, which does not meet the age criteria. Additionally, there is no evidence of cirrhosis in this patient. Therefore, HCC surveillance may not be warranted at this time. | No | No - 43.4% |
| ***A 25-year-old Vietnamese man with past history of hepatitis B (hepatitis B surface antigen negative, hepatitis B core antibody positive,*** | Based on the provided context, this patient does not need to be screened for hepatocellular carcinoma. The patient is a young man from an endemic country (Vietnam), but he is under the age of 40. Additionally, his hepatitis B surface antigen (HBsAg) is negative, which indicates that he is not currently infected with hepatitis B. Therefore, he does not meet the criteria for | No | No - 46.7% |
| **hepatitis B surface antibody positive).** | hepatocellular carcinoma screening according to the provided guidelines. |  |  |
| **A 35-year-old Chinese woman with platelets of 80,000, INR of 1.1, bilirubin of 1.4 mg/dL. On ultrasound, she was noted to have mild ascites and a spleen that measures 14 cm.** | Based on the provided context, the patient should be considered for hepatocellular carcinoma (HCC) screening. The presence of ascites and a low platelet count may suggest advanced liver disease or cirrhosis. Regular screening for HCC with cross-sectional imaging at 6-month intervals is advised for patients with cirrhosis. However, the final decision should be made by a healthcare professional considering the patient's overall health status and other risk factors. | Yes | Yes - 76.8% |
| **A 40-year-old woman from Laos with chronic inactive hepatitis B without cirrhosis.** | Based on the provided information, a 40-year-old woman from Laos (an endemic country as defined by AASLD hepatitis B virus guidance) with chronic inactive hepatitis B without cirrhosis does not meet the criteria for sufficient risk to warrant surveillance for hepatocellular carcinoma (HCC). The guidelines suggest surveillance for women from endemic countries when they are over the age of 50. | No | No - 34.8% |
| **A 40-year-old woman from Thailand with cirrhosis and chronic inactive hepatitis B. \*** | Yes, the patient should be screened for hepatocellular carcinoma. According to the provided context, individuals with Child-Pugh A –B cirrhosis, any etiology, and chronic hepatitis B are at sufficient risk to warrant surveillance for hepatocellular carcinoma. Additionally, the patient is from an endemic country (as defined by AASLD hepatitis B virus guidance) and is over the age of 40, which further supports the need for screening. | Yes | Yes - 81.2% |
| **A 40-year-old Nigerian woman with chronic hepatitis B with normal liver function tests.** | Yes, this patient should be screened for hepatocellular carcinoma. The guidelines suggest that a person from Africa, regardless of gender, should start surveillance as early as the third decade of life given the median age of 46 years at HCC diagnosis. This is especially relevant if the person has chronic hepatitis B, which is a known risk factor for hepatocellular carcinoma. | Yes | Yes - 65.8% |
| **\*LiVersa's generated answer and rationale may not be completely correct in these cases.** |  |  |  |

## Discussion

Two of the most significant barriers to using general-purpose LLMs like ChatGPT in clinical practice are the tendency for these LLMs to “hallucinate” or generate confident sounding but false responses, and their lack of compatibility with PHI.^6,7^ The first issue stems from the nature of the transformer architecture,^25^ which optimizes the LLM’s objective of predicting the most probable next word (token) from its pre-trained data without consideration for accuracy. Augmenting the knowledge base of general-purpose LLMs, such as through RAG, becomes one of the key strategies to shaping and constraining LLM outputs to prevent false information from being propagated and disseminated. Our work is one of the first demonstrations and proof-of-concept of using RAG to create a liver disease-specific and, more importantly, PHI-compliant LLM chat interface. Our incorporation of hepatology specific knowledge, such as the AASLD practice guidelines, resulted in the LiVersa chat interface generating answers that were likely more specific than those generated by general-purpose ChatGPT.^24^

While LiVersa answered all 10 questions regarding HBV treatment and HCC surveillance in our test set correctly, the rationales given for three cases were not completely correct. These incorrect responses reflect limitations of utilizing RAG. For instance, LiVersa’s stated rationale for a “Yes” recommendation for the case-vignette of “A 25-year-old Haitian man with chronic hepatitis B, on treatment with entecavir” was based on the presumption that the patient was from an HBV endemic country (Table 3). The actual rationale given in prior literature is based on the 2018 version of the AASLD HCC practice guidance document, which recommended early surveillance for African HBV patients and/or North American HBV patients of African-American descent.

In this example, we highlight two key issues. First, the 2018 version of the AASLD HCC practice guidance document was not uploaded into LiVersa’s RAG dataset, the 2023 version was.^26,27^ LiVersa gave the wrong justification for HCC surveillance in this case simply because the 2018 version was not available to it and “restrictions” placed by the RAG dataset forced it to generate a hallucinatory answer. This limitation could easily be overcome by increasing the number and variety of literature incorporated into LiVersa’s RAG dataset. Additional documents that could be considered for inclusion in the future include clinical practice guidelines/guidance documents from other societies, such as those from the American Gastroenterological Association, American College of Gastroenterology, European Association for the Study of the Liver, and Asian Pacific Association for the Study of the Liver; and from compendium sources, such as the Cochrane Review and UpToDate.

The second key issue is contextual bias. In this instance, the recommendation for HCC surveillance is based on the assumption that the man from Haiti in the clinical scenario is of African descent. This is a problem that affects both literature data incorporated into the RAG dataset and in the pre-training data that underpins the LLM itself. The more “correct” response should have been to ask for additional context to clarify the self-identified national-origin or racial/ethnic background of the patient to allow the LLM to give a more comprehensive recommendation/output. This is an illustration of the problems concerning algorithm bias that generative artificial intelligence (GAI) research community is actively grappling with.^28–30^ Finally, in addition to the two key considerations noted above, there is a minor limitation with regards to our grading of LiVersa’s responses. We had edited the case-vignette knowledge assessment questions for clarity and appropriateness. While we tried to retain the intent of the clinical cases, the wording of these cases may not be the same as those evaluated by trainees and ChatGPT in prior studies.^23,24^ This may have affected the comparisons in Table 3.

Despite these limitations, we demonstrated that RAG could be a powerful method to create a “specialized” LLM. Given that our LiVersa prototype was developed and deployed in a PHI-compliant environment (UCSF’s Versa), it could theoretically be used clinically to evaluate actual patient scenarios. By extension, there is a potential to incorporate both literature and patient data from the electronic health record through extraction by the Fast Healthcare Interoperability Resources (FHIR) API in the RAG process.^31^ This process would allow for the creation of patient-specific clinical LLMs that could be a true realization of GAI-enabled personalized medicine.

## Data Availability

The clinical guidelines/guidance documents used in the methods of this manuscript are publicly available.

## Acknowledgements

- The authors thank the UCSF AI Tiger Team, Academic Research Services, Research Information Technology, and the Chancellor’s Task Force for Generative AI for their software development, analytical, and technical support related to the use of Versa API gateway (the UCSF secure implementation of large language models and generative AI via API gateway), Versa chat (the chat user interface), and related data assets.

## Abbreviations

AASLD: American Association for the Study of Liver Diseases
APIs: application programming interfaces
FHIR: Fast Healthcare Interoperability Resources
HBV: hepatitis B
HCC: hepatocellular carcinoma
GAI: generative artificial intelligence
GPT: generative pre-trained transformer
LLMs: large language models
PHI: protected health information
RAG: retrieval-augmented generation
UCSF: University of California, San Francisco

## Writing Assistance

None.

## Author Contributions

Authorship was determined using ICMJE recommendations.

*Ge:* Concept and design; data extraction; analysis and interpretation of data; drafting of manuscript; critical revision of the manuscript for important intellectual content; study supervision

*Sun:* Data extraction; analysis and interpretation of data; critical revision of the manuscript for important intellectual content

*Owens:* Concept and design; analysis and interpretation of data; critical revision of the manuscript for important intellectual content

*Galvez:* Analysis and interpretation of data; critical revision of the manuscript for important intellectual content

*Gologorskaya:* Analysis and interpretation of data; critical revision of the manuscript for important intellectual content

*J. Lai:* Concept and design; critical revision of the manuscript for important intellectual content; study supervision

*Pletcher:* Concept and design; critical revision of the manuscript for important intellectual content; study supervision

*K. Lai:* Concept and design; critical revision of the manuscript for important intellectual content; study supervision

## References

1. Ge J, Li M, Delk MB, Lai JC. A comparison of large language model versus manual chart review for extraction of data elements from the electronic health record. medRxiv. September 1, 2023.

2. Rahman M, Terano HJR, Rahman N, Salamzadeh A, Rahaman S. Chatgpt and academic research: A review and recommendations based on practical examples. J Educ, Mngt, and Dev Studies. 2023;3(1):1–12.

3. Nayak A, Alkaitis MS, Nayak K, Nikolov M, Weinfurt KP, Schulman K. Comparison of history of present illness summaries generated by a chatbot and senior internal medicine residents. JAMA Intern Med. 2023;183(9):1026–1027.

4. Han C, Kim DW, Kim S, et al. Evaluation Of GPT-4 for 10-Year Cardiovascular Risk Prediction: Insights from the UK Biobank and KoGES Data. 2023.

5. ChatGPT: Optimizing Language Models for Dialogue. Accessed December 17, 2022. https://openai.com/blog/chatgpt/

6. Ge J, Lai JC. Artificial intelligence-based text generators in hepatology: ChatGPT is just the beginning. Hepatol Commun. 2023;7(4).

7. Ji Z, Lee N, Frieske R, et al. Survey of hallucination in natural language generation. ACM Comput Surv. November 17, 2022.

8. Densen P. Challenges and opportunities facing medical education. Trans Am Clin Climatol Assoc. 2011;122:48–58.

9. Landhuis E. Scientific literature: Information overload. Nature. 2016;535(7612):457-458.

10. Practice Guidelines | AASLD. Accessed November 8, 2023. https://www.aasld.org/practice-guidelines

11. GPT-3.5 Turbo fine-tuning and API updates. Accessed November 8, 2023. https://openai.com/blog/gpt-3-5-turbo-fine-tuning-and-api-updates

12. Jiang LY, Liu XC, Nejatian NP, et al. Health system-scale language models are all-purpose prediction engines. Nature. 2023;619(7969):357–362.

13. Kojima T, Gu SS, Reid M, Matsuo Y, Iwasawa Y. Large Language Models are Zero-Shot Reasoners. arXiv. 2022.

14. Brown TB, Mann B, Ryder N, et al. Language models are few-shot learners. arXiv. 2020.

15. Parnami A, Lee M. Learning from Few Examples: A Summary of Approaches to Few-Shot Learning. arXiv. 2022.

16. RAG and generative AI - Azure Cognitive Search | Microsoft Learn. Accessed November 8, 2023. https://learn.microsoft.com/en-us/azure/search/retrieval-augmented-generation-overview

17. Wang Y, Ma X, Chen W. Augmenting Black-box LLMs with Medical Textbooks for Clinical Question Answering. arXiv. 2023.

18. Lozano A, Fleming SL, Chiang C-C, Shah N. Clinfo.ai: An Open-Source Retrieval-Augmented Large Language Model System for Answering Medical Questions using Scientific Literature. arXiv. 2023.

19. Khene Z-E, Bigot P, Mathieu R, Rouprêt M, Bensalah K, French Committee of Urologic Oncology. Development of a personalized chat model based on the european association of urology oncology guidelines: harnessing the power of generative artificial intelligence in clinical practice. Eur Urol Oncol. July 18, 2023.

20. Ferber D, Kather JN. Large Language Models in Uro-oncology. Eur Urol Oncol. October 13, 2023.

21. Embeddings - OpenAI API. Accessed October 27, 2023. https://platform.openai.com/docs/guides/embeddings

22. New and improved embedding model. Accessed October 27, 2023. https://openai.com/blog/new-and-improved-embedding-model

23. Mahfouz M, Nguyen H, Tu J, et al. Knowledge and perceptions of hepatitis B and hepatocellular carcinoma screening guidelines among trainees: A tale of three centers. Dig Dis Sci. 2020;65(9):2551–2561.

24. Yeo YH, Samaan JS, Ng WH, et al. Assessing the performance of ChatGPT in answering questions regarding cirrhosis and hepatocellular carcinoma. Clin Mol Hepatol. 2023;29(3):721–732.

25. Vaswani A, Shazeer N, Parmar N, et al. Attention is all you need. arXiv. 2017.

26. Marrero JA, Kulik LM, Sirlin CB, et al. Diagnosis, staging, and management of hepatocellular carcinoma: 2018 practice guidance by the american association for the study of liver diseases. Hepatology. 2018;68(2):723–750.

27. Singal AG, Llovet JM, Yarchoan M, et al. AASLD Practice Guidance on prevention, diagnosis, and treatment of hepatocellular carcinoma. Hepatology. May 22, 2023.

28. Fang X, Che S, Mao M, Zhang H, Zhao M, Zhao X. [2309.09825] Bias of AI-Generated Content: An Examination of News Produced by Large Language Models. arXiv. September 18, 2023.

29. Zack T, Lehman E, Suzgun M, et al. Coding Inequity: Assessing GPT-4’s Potential for Perpetuating Racial and Gender Biases in Healthcare. medRxiv. July 16, 2023.

30. DeCamp M, Lindvall C. Latent bias and the implementation of artificial intelligence in medicine. J Am Med Inform Assoc. 2020;27(12):2020–2023.

31. Braunstein ML. Pre-FHIR Interoperability and Clinical Decision Support Standards. In: Health Informatics on FHIR: How Hl7’s New API Is Transforming Healthcare. Springer International Publishing; 2018:151–177.

32. Lee WM, Larson AM, Stravitz RT. AASLD position paper: the management of acute liver failure: update 2011. Hepatology. 2011;55(3):965–967.

33. Crabb DW, Im GY, Szabo G, Mellinger JL, Lucey MR. Diagnosis and Treatment of Alcohol-Associated Liver Diseases: 2019 Practice Guidance From the American Association for the Study of Liver Diseases. Hepatology. 2020;71(1):306–333.

34. Biggins SW, Angeli P, Garcia-Tsao G, et al. Diagnosis, evaluation, and management of ascites, spontaneous bacterial peritonitis and hepatorenal syndrome: 2021 practice guidance by the american association for the study of liver diseases. Hepatology. 2021;74(2):1014–1048.

35. Mack CL, Adams D, Assis DN, et al. Diagnosis and management of autoimmune hepatitis in adults and children: 2019 practice guidance and guidelines from the american association for the study of liver diseases. Hepatology. 2020;72(2):671–722.

36. Fontana RJ, Liou I, Reuben A, et al. AASLD practice guidance on drug, herbal, and dietary supplement-induced liver injury. Hepatology. 2023;77(3):1036–1065.

37. Bacon BR, Adams PC, Kowdley KV, Powell LW, Tavill AS, American Association for the Study of Liver Diseases. Diagnosis and management of hemochromatosis: 2011 practice guideline by the American Association for the Study of Liver Diseases. Hepatology. 2011;54(1):328–343.

38. Vilstrup H, Amodio P, Bajaj J, et al. Hepatic encephalopathy in chronic liver disease: 2014 Practice Guideline by the American Association for the Study of Liver Diseases and the European Association for the Study of the Liver. Hepatology. 2014;60(2):715–735.

39. Terrault NA, Bzowej NH, Chang K-M, et al. AASLD guidelines for treatment of chronic hepatitis B. Hepatology. 2016;63(1):261–283.

40. Terrault NA, Lok ASF, McMahon BJ, et al. Update on prevention, diagnosis, and treatment of chronic hepatitis B: AASLD 2018 hepatitis B guidance. Hepatology. 2018;67(4):1560–1599.

41. Bhattacharya D, Aronsohn A, Price J, Lo Re V, AASLD-IDSA HCV Guidance Panel. Hepatitis C Guidance 2023 Update: AASLD-IDSA Recommendations for Testing, Managing, and Treating Hepatitis C Virus Infection. Clin Infect Dis. May 25, 2023.

42. Rockey DC, Caldwell SH, Goodman ZD, Nelson RC, Smith AD, American Association for the Study of Liver Diseases. Liver biopsy. Hepatology. 2009;49(3):1017–1044.

43. Martin P, DiMartini A, Feng S, Brown R, Fallon M. Evaluation for liver transplantation in adults: 2013 practice guideline by the American Association for the Study of Liver Diseases and the American Society of Transplantation. Hepatology. 2014;59(3):1144–1165.

44. Squires RH, Ng V, Romero R, et al. Evaluation of the pediatric patient for liver transplantation: 2014 practice guideline by the American Association for the Study of Liver Diseases, American Society of Transplantation and the North American Society for Pediatric Gastroenterology, Hepatology and Nutrition. Hepatology. 2014;60(1):362–398.

45. Lucey MR, Terrault N, Ojo L, et al. Long-term management of the successful adult liver transplant: 2012 practice guideline by the American Association for the Study of Liver Diseases and the American Society of Transplantation. Liver Transpl. 2013;19(1):3–26.

46. Kelly DA, Bucuvalas JC, Alonso EM, et al. Long-term medical management of the pediatric patient after liver transplantation: 2013 practice guideline by the American Association for the Study of Liver Diseases and the American Society of Transplantation. Liver Transpl. 2013;19(8):798–825.

47. Lai JC, Tandon P, Bernal W, et al. Malnutrition, frailty, and sarcopenia in patients with cirrhosis: 2021 practice guidance by the american association for the study of liver diseases. Hepatology. 2021;74(3):1611–1644.

48. Rinella ME, Neuschwander-Tetri BA, Siddiqui MS, et al. AASLD Practice Guidance on the clinical assessment and management of nonalcoholic fatty liver disease. Hepatology. 2023;77(5):1797–1835.

49. Rogal SS, Hansen L, Patel A, et al. AASLD Practice Guidance: Palliative care and symptom-based management in decompensated cirrhosis. Hepatology. 2022;76(3):819–853.

50. Kaplan DE, Bosch J, Ripoll C, et al. AASLD practice guidance on risk stratification and management of portal hypertension and varices in cirrhosis. Hepatology. October 23, 2023.

51. Lee EW, Eghtesad B, Garcia-Tsao G, et al. AASLD practice guidance on the use of TIPS, variceal embolization, and retrograde transvenous obliteration in the management of variceal hemorrhage. Hepatology. June 30, 2023.

52. Lindor KD, Bowlus CL, Boyer J, Levy C, Mayo M. Primary Biliary Cholangitis: 2018 Practice Guidance from the American Association for the Study of Liver Diseases. Hepatology. 2019;69(1):394–419.

53. Lindor KD, Bowlus CL, Boyer J, Levy C, Mayo M. Primary biliary cholangitis: 2021 practice guidance update from the American Association for the Study of Liver Diseases. Hepatology. 2022;75(4):1012–1013.

54. Bowlus CL, Arrivé L, Bergquist A, et al. AASLD practice guidance on primary sclerosing cholangitis and cholangiocarcinoma. Hepatology. 2023;77(2):659–702.

55. Sarkar M, Brady CW, Fleckenstein J, et al. Reproductive health and liver disease: practice guidance by the american association for the study of liver diseases. Hepatology. 2021;73(1):318–365.

56. Northup PG, Garcia-Pagan JC, Garcia-Tsao G, et al. Vascular liver disorders, portal vein thrombosis, and procedural bleeding in patients with liver disease: 2020 practice guidance by the american association for the study of liver diseases. Hepatology. 2021;73(1):366–413.

57. Schilsky ML, Roberts EA, Bronstein JM, et al. A multidisciplinary approach to the diagnosis and management of Wilson disease: 2022 Practice Guidance on Wilson disease from the American Association for the Study of Liver Diseases. Hepatology. December 7, 2022.

58. Kanwal F, Tapper EB, Ho C, et al. Development of quality measures in cirrhosis by the practice metrics committee of the american association for the study of liver diseases. Hepatology. 2019;69(4):1787–1797.

59. Asrani SK, Ghabril MS, Kuo A, et al. Quality measures in HCC care by the Practice Metrics Committee of the American Association for the Study of Liver Diseases. Hepatology. 2022;75(5):1289–1299.

60. Karvellas CJ, Bajaj JS, Kamath PS, et al. AASLD Practice guidance on Acute-on-chronic liver failure and the management of critically Ill patients with cirrhosis. Hepatology. November 9, 2023.

